# Prediction of the need for intensive oxygen supplementation during the hospitalization among patients with COVID 19 admitted to an academic health system in Texas, USA: a retrospective cohort study and development of a multivariable regression model

**DOI:** 10.1101/2021.11.05.21265970

**Authors:** John W. Davis, Beilin Wang, Ewa Tomczak, Chia-Chi Fu, Wissam Harmouch, David Reynoso, Philip Keiser, Miguel M. Cabada

## Abstract

**Objective:** The severe acute respiratory syndrome-Coronavirus-2 (SARS-CoV-2) has caused a pandemic claiming more than 4 million lives worldwide. Overwhelming Coronavirus-Disease-2019 (COVID-19) respiratory failure placed tremendous demands on healthcare systems increasing the death toll. Cost-effective prognostic tools to characterize COVID-19 patients’ likely to progress to severe hypoxemic respiratory failure are still needed.

**Design:** We conducted a retrospective cohort study to develop a model utilizing demographic and clinical data collected in the first 12-hours admission to explore associations with severe hypoxemic respiratory failure in unvaccinated and hospitalized COVID-19 patients.

**Setting:** University based healthcare system including 6 hospitals located in the Galveston, Brazoria and Harris counties of Texas.

**Participants:** Adult patients diagnosed with COVID-19 and admitted to one of six hospitals between March 19^th^ and June 31^st^, 2020.

**Primary outcome:** The primary outcome was defined as reaching a WHO ordinal scale between 6-9 at any time during admission, which corresponded to severe hypoxemic respiratory failure requiring high-flow oxygen supplementation or mechanical ventilation.

**Results:** We included 329 participants in the model cohort and 62 (18.8%) met the primary outcome. Our multivariable regression model found that lactate dehydrogenase (OR 2.36), qSOFA score (OR: 2.26), and neutrophil to lymphocyte ratio (OR:1.15) were significant predictors of severe disease. The final model showed an area under curve (AUC) of 0.84. The sensitivity analysis and point of influence analysis did not reveal inconsistencies.

**Conclusions:** Our study suggests that a combination of accessible demographic and clinical information collected on admission may predict the progression to severe COVID-19 among adult patients with mild and moderate disease. This model requires external validation prior to its use.

**STRENGTHS AND LIMITATIONS OF THIS STUDY:**

⍰ Our study utilized objective and measurable demographic and clinical information regularly available in healthcare settings even among patients unable to communicate.
⍰ Our primary outcome corresponds to WHO ordinal score which would allow compare our results to other studies and in other settings.
⍰ Our model could serve as an effective point of service tool during early admission to assist in clinical management and allocation of resources to unvaccinated patients.
⍰ Our study is a retrospective study of unvaccinated COVID19 patients, and validation of our prediction model in the rest of our study population is still needed.
⍰ In addition, testing our model in a more recent cohort after emergence of new SARS-CoV-2 variants will be needed to assess its robustness.

## INTRODUCTION

The severe acute respiratory syndrome-Coronavirus-2 (SARS-CoV-2) is a novel coronavirus discovered in 2019. It is the etiologic agent for the largest viral pandemic of the 21^st^ century thus far, followed by H1N1 Influenza A emerged in 2009-2010..[1, 2] During the early pandemic, a case series from the Wuhan province showed that 81% of COVID-19 cases were mild, 14% progressed to severe disease, and 5% developed critical illness defined as respiratory failure, septic shock and/or multiple organ dysfunction..[3] COVID19 associated hospitalizations caused an overwhelming demand on the healthcare system of the United States. Shortage in ventilators and personal protection equipment posed significant challenges in management of cases in US hospitals early in the pandemic.[13] During 2020, CDC estimated 345,000 deaths attributed to COVID19.[14]

Unfortunately, there has been a global lag in uptake of COVID-19 vaccines due to hesitancy and logistics. Unvaccinated COVID-19 individuals remain up to 25 times more likely to be hospitalized or dead compared to vaccinated individuals. Rising hospitalizations and deaths among unvaccinated individuals are driving a new pandemic surge posing again a significant burden to the health system.[19] Studies evaluating the risk of progression among infected subjects admitted to the hospital have used different outcomes to define severe diseases. These included criteria from the American Thoracic Society’s on severity of community acquired pneumonia [4], the Berlin definition of acute respiratory distress syndrome [5], death or mechanical ventilation [6, 7], and/or the World Health Organization (WHO) ordinal scale.[8, 9] The WHO ordinal scale to classify the clinical status of patients with COVID-19 has been widely adopted in randomized control trials such as ACTT-1 and ACTT-2.[10-12] Harmonization of the measures used to evaluate the severity COVID-19 across different studies could ease the comparison of study results and application of evidence-based interventions. However, the heterogeneity in the definitions of severe illness and the limited availability of certain laboratory tests, especially in low-resource settings, have decreased the generalizability of these tools. Laboratory tests such as serum IL-6 or procalcitonin may not be accessible in small medical centers. Similarly, information on comorbidities may not be available in patients unable to provide a history. Simple, objective, and accessible tools to predict progression to severe COVID 19 are still needed to guide clinicians during case surges and dwindling of resources. To address this need, we conducted a retrospective cohort study in the University of Texas Medical Branch Health System to develop an exploratory model for severe hypoxemic respiratory failure in unvaccinated, hospitalized COVID-19 patients.

## METHODS

### Study Design

We hypothesized that a combination of objective clinical and laboratory findings on admission can identify subjects with higher risk of progression to severe respiratory failure due to COVID-19 in our hospitals. To test this hypothesis, we performed a retrospective, multi-site cohort study on adult patients admitted for COVID-19 to the University of Texas Medical Branch (UTMB) Health System.

UTMB’s health system includes 6 hospitals located in the Galveston, Brazoria, and Harris counties of Texas. These hospitals are distributed across over 50 miles, though populations served are similar overall. We retrieved the medical record numbers of all patients ≥ 18 years old admitted to hospitals in any of the four campuses with a positive SARS-CoV-2 molecular test between March 19^th^ and June 31^st^ of 2020. We used the WHO ordinal scale of disease severity for COVID-19 to define our outcomes.[25] This is an eleven-category ordinal scale ranging from a value of zero for patients with no virological evidence of infection to 10 for patients who died due to COVID-19. Our primary outcome was defined as reaching a WHO ordinal scale between 6-9 during admission corresponding to severe respiratory failure requiring oxygen supplementation using HFNC or mechanical ventilation. Patients initially presenting with a WHO ordinal scale <6 that were discharged at the time of review of their medical record were enrolled. Patients who met ordinal scale 6-9 on the first vital signs obtained on admission were excluded. The maximum ordinal scale score met during admission was considered the subjects’ ordinal score. The study protocol was approved by UTMB Institutional Review Board (20-0126) and the Texas Department of Criminal Justice (TDCJ) Institutional Review Board (#819-RM20).

### Patient or Public Involvement

None.

### Data Collection

We collected data directly from the Epic (Verona, Wisconsin) electronic medical records. The data was transcribed into a questionnaire created in the REDCap (Nashville, Tennessee) data capture system. Data coders were trained using a dummy dataset before using medical records. All coders were trained until they could obtain 100% accuracy on dummy datasets before proceeding to data collection. Eighty-nine randomly selected charts underwent evaluation by the principal investigators and the data extraction personnel. These evaluations were compared to calculate the inter-rater reliability using Kappa statistics. When the personnel had a Kappa < 0.8, they were re-trained, and discrepancies were discussed with the principal investigators. Evaluations were repeated until a Kappa >0.8 was reached. The data extraction personnel collected data on demographics, clinical history and course, vital signs, peak oxygen requirement, and laboratory results (Supplemental eTable A). The maximum oxygen requirement at any given day after admission was used as the peak oxygen requirement, and the subject was deemed to have met the primary endpoint if the peak oxygen requirements was high flow nasal canula or more intensive. Data on admission laboratory results include absolute neutrophil and lymphocyte counts; serum lactate dehydrogenase (LDH), D-dimer, C-reactive protein (CRP), procalcitonin, and troponin I. Only the first laboratory tests obtained within 12 hours of admission were recorded. If these labs were not obtained during this window, they were registered as missing.

### Statistical Analysis

The REDCap dataset was downloaded to a database on SAS (Version 9.4, Cary NC) and R (Version 4.0.2). Frequencies, means with standard deviations (±SD), and medians with interquartile ranges (IQR) were calculated to describe the distribution of the variables. Pearson correlations were performed for bivariate analysis; variance inflation factors (VIFs) were calculated for each variable prior to initial modeling, with a factor >= 5 being considered possibly collinear.[13] Mean imputation was used to replace BMI when the value was missing. Multiple imputation was not performed because data were not missing at random relative to the primary outcome. To evaluate the effect of utilizing the sample mean to replace BMI missing values, the analysis was also performed excluding those cases.

A multivariable logistic regression analysis was conducted to model variables with the highest predictive value for severe COVID-19. Variables for the model were selected based on review of the literature on COVID-19 and clinical relevance (e.g, objectivity and availability). The composite variables age/BMI and age/sex were created and utilized in the models because of existing evidence of interactions between those variables individually. Stratified analysis was performed for interaction terms ultimately included in the model. Stepwise Akaike Information Criteria (AIC) reduction was utilized to optimize the model, reducing residual deviance while prioritizing model simplicity. Cook’s Distance method was utilized for assessing points of influence, where a Cook’s D ≥ 1 was considered highly influential. The Hoslem-Lemeshow goodness of fit (GOF) test statistic was utilized to evaluate the match between the predicted and observed risk of progression to WHO ordinal score 6 to 9. A receiver operating characteristics curve (ROC) and area under the curve (AUC) analysis was performed to assess overall model fidelity.

Several sensitivity analyses were performed. One was to assess the biasing effect of mean imputation on BMI. We excluded all cases where BMI was missing for this analysis. The second was to assess whether DNI status meaningfully affected results. Because some patients may have initiated DNI during the course of admission (which we could not verify), we excluded all patients who had a DNI in place by the time of discharge or death. The final was to examine whether a model that ordinally discriminated between HFNC and intubation was more robust (where < HFNC=3, HFNC=5, and Intubation=6; values according to WHO Scale). Because comparative AUC analysis between cumulative logit and binomial logistic regression is not possible, qualitative differences parameter selection, magnitude, and percent concordance were assessed. Proportional odds assumptions were tested using Chi Square methods.

## RESULTS

We identified 930 subjects admitted to the UTMB Health System with a positive SARS-CoV-2 test during March 19^th^ – June 31^st^, 2020. The first 352 consecutive charts were reviewed to develop the predictive models. The demographics and clinical characteristics of the cohort prior to exclusion are shown in Supplemental Figure 1. 23 subjects were excluded because they met WHO ordinal scale scores between 6-9 on the first vital signs measured or because most values of interest were missing (Figure 1). 329 subjects were included in the final cohort and 62 (18.8%) met the primary endpoint. The TDCJ population accounted for 27.6% of cohort population but there were no significant differences in the proportion of subjects meeting the primary endpoint according to inmate status (p=0.459, data not shown). Subjects reaching the ordinal scale 6-9 were significantly older than subjects who did not (Table 1). More male subjects met the primary end point but the difference between groups was not statistically significant (Table 1). The top three comorbidities for subjects with ordinal scales <6 were cardiovascular 51.5%, diabetes mellitus 32.4%, and pulmonary 21.0%. For subjects with ordinal scale 6-9, the top three conditions were cardiovascular 67.3%, diabetes mellitus 38.5%, and liver disease 15.4% (Table 1). 24 subjects died during admission (7.3%) and 20 of them met criteria for ordinal score 6-9. 6.7% of subjects with ordinal score <6 and 9.7% of subjects with ordinal scale 6-9 had DNI order. Comfort care was implemented in 1.9% of those with ordinal score <6 and in 16.1% of those with ordinal score 6-9. The characteristics of subjects with ordinal scales 6-9 across all campuses are shown (Table 1). 14 (4.3%) were missing BMI values.

**Figure 1:**
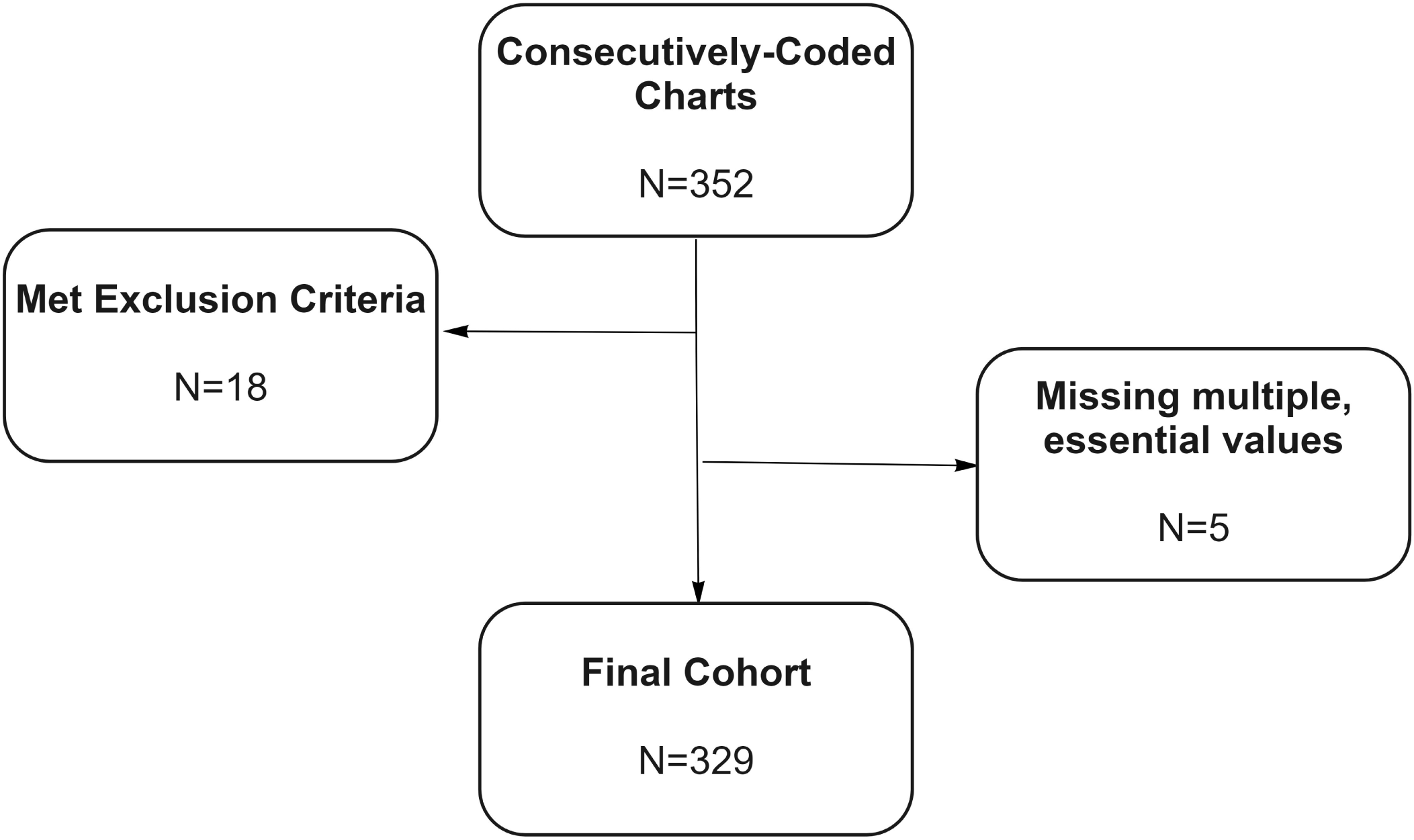
Flow Chart for Cohort Selection

**Table 1.**
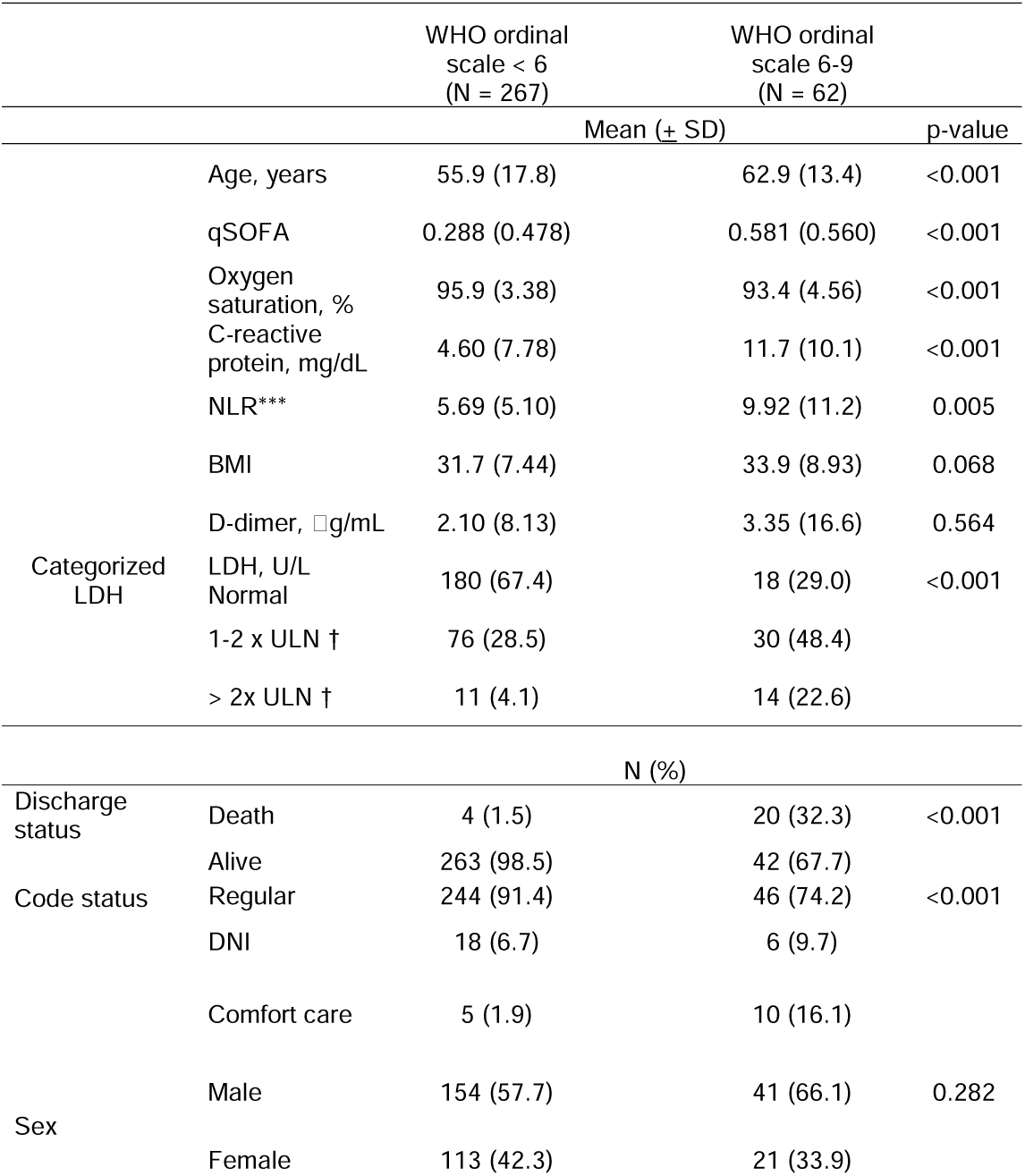
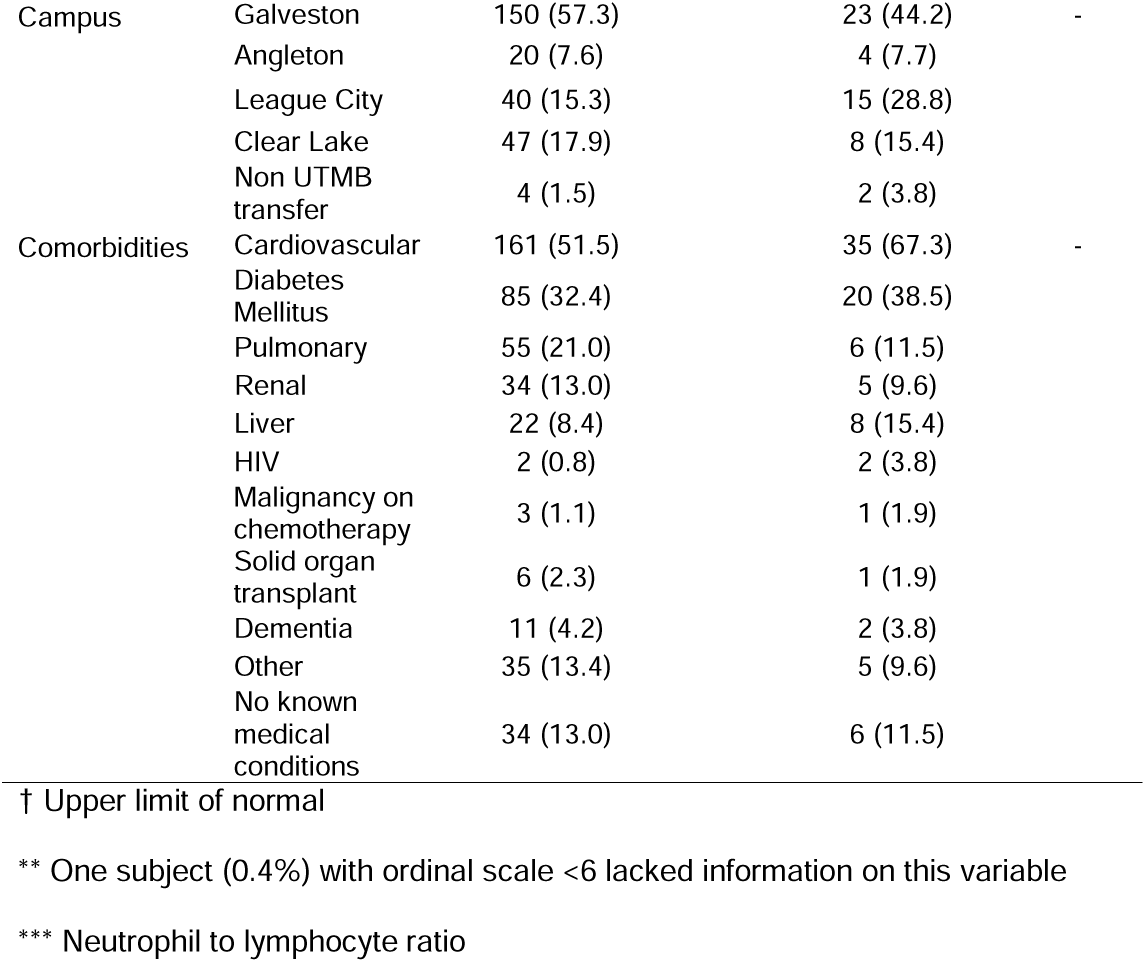
Demographic and clinical characteristics of subjects included in the cohort.

The variables included in the initial regression model were admission date, age/sex, age/BMI, oxygen saturation, neutrophil to lymphocyte ratio (NLR), procalcitonin, D-dimer, LDH, CRP, troponin I, duration of symptoms prior to admission, and qSOFA score (Supplemental Figure 2).

The initial model was highly significant and identified several candidate predictive variables; none of these variables had a VIF >3. The candidate clinical and laboratory variables age, BMI, oxygen saturation, qSOFA score, CRP, procalcitonin, NLR, D-dimer and LDH were incorporated into prognostic model. All subjects with elevated troponin I levels (6/6) were intubated which precluded the evaluation of this variable as a predictor in the analysis. After stepwise AIC reduction, the final model included 7 variables: oxygen saturation, NLR, D-dimer, qSOFA, LDH, age, BMI, and admission date (Table 2). Because admission date did not meaningfully improve model performance (AUC=0.84 without vs 0.85 with) and complicates clinical use, we exclude that factor here.

**Table 2.**
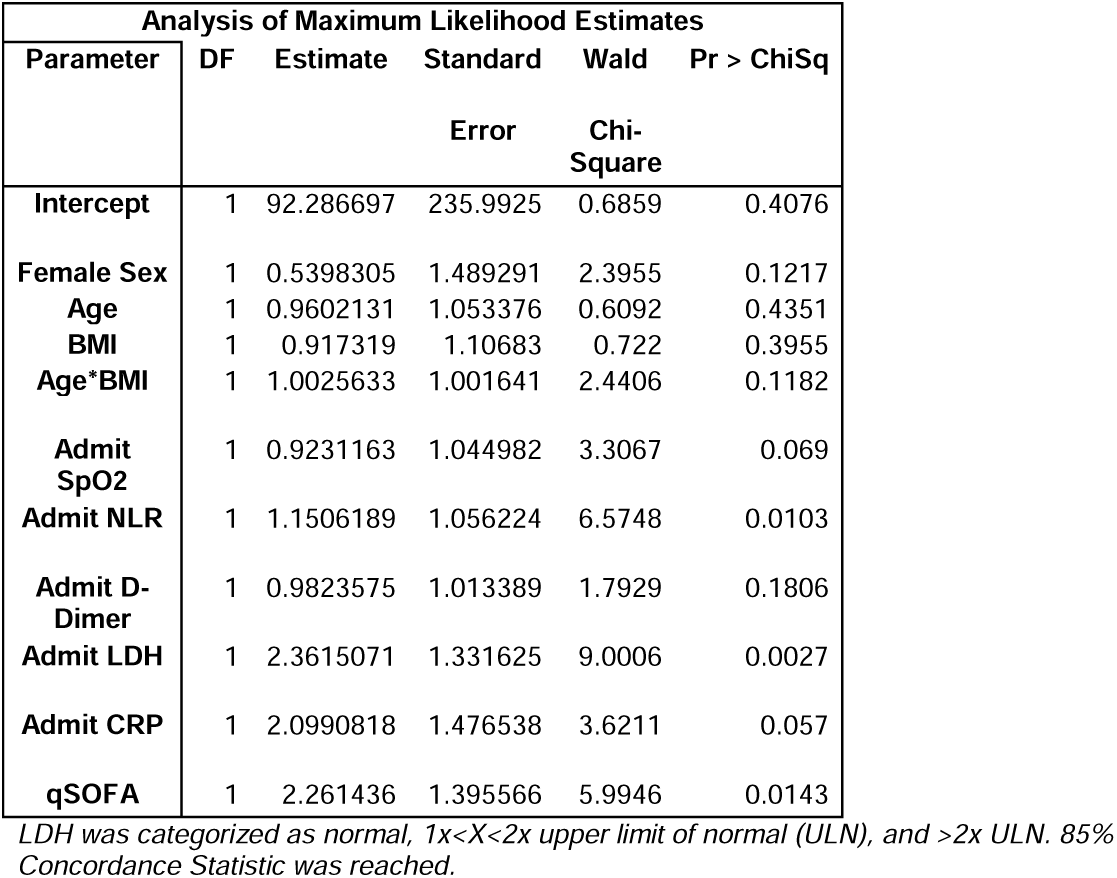
Final multivariable regression model after stepwise AIC reduction.

Stratified analysis (BMI >= 30 vs BMI<30) was limited due to small sample of patients with lower BMIs. The higher BMI strata, however, demonstrated similar effect sizes for age and BMI; qSOFA score was not retained (Supplemental Figure 3). Running the model excluding subjects with missing BMI values (n=34) did not affect the general significance or goodness of fit of the model (Table 3).

**Table 3.**
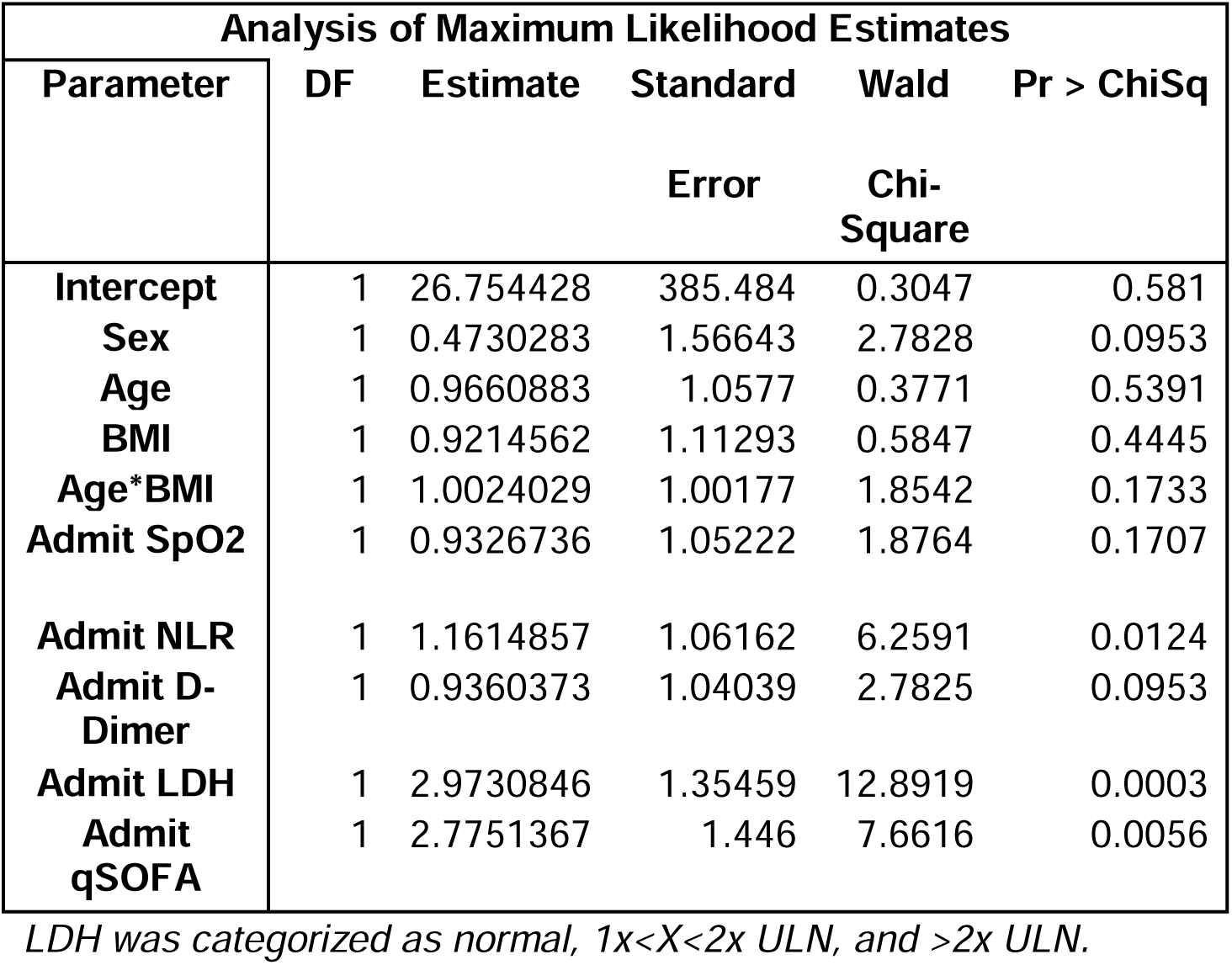
Multiple regression model excluding cases with missing BMI.

ROC/AUC analysis of the final model indicated an area under the curve of 0.84, indicating high efficacy of the overall model in predicting severe disease (Figure 2).

**Figure 2:**
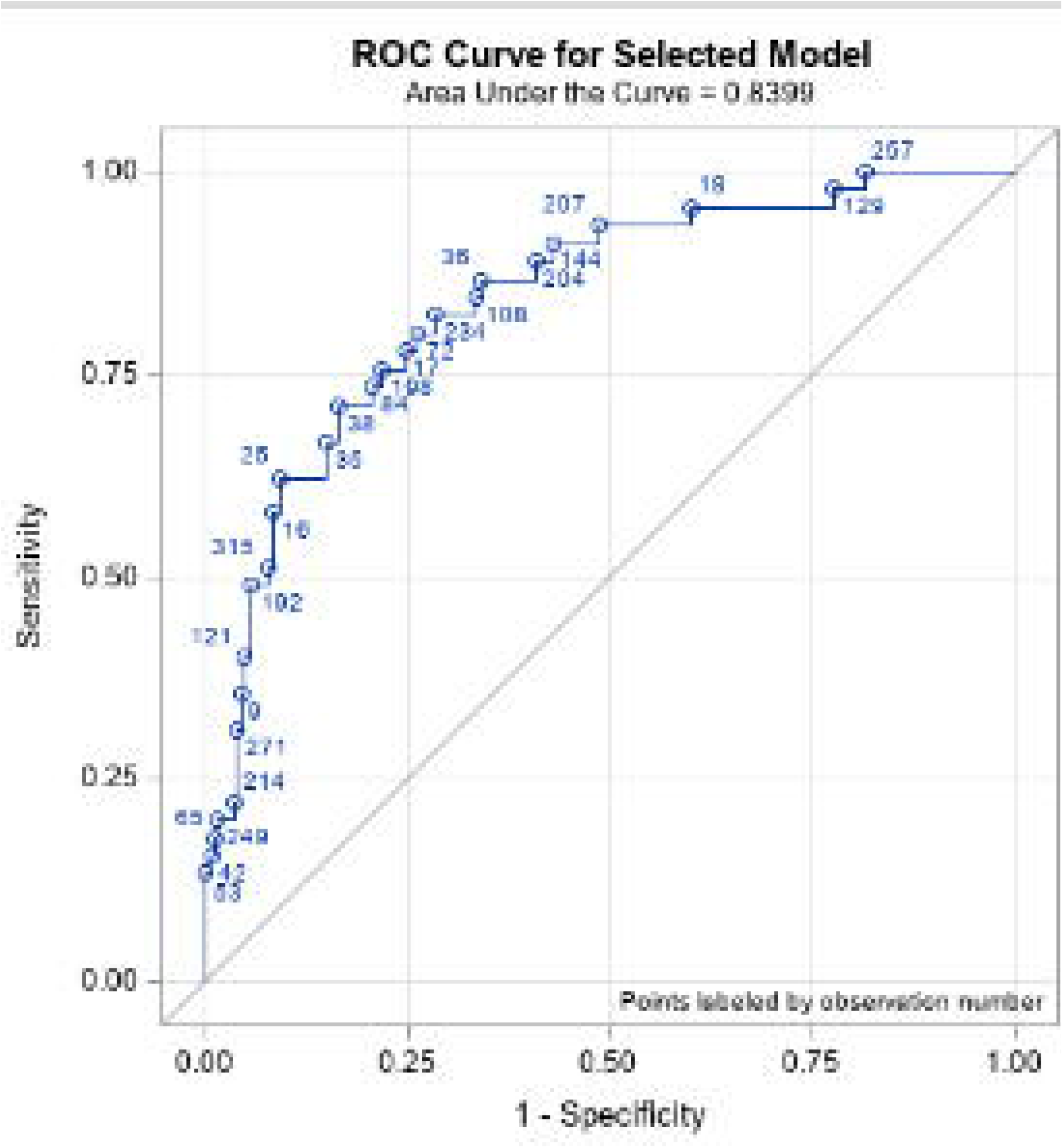
ROC Curve, Final Model

ROC/AUC analysis of the BMI sensitivity analysis was nearly identical to the final model (AUC = 0.83; Figure 3).

**Figure 3:**
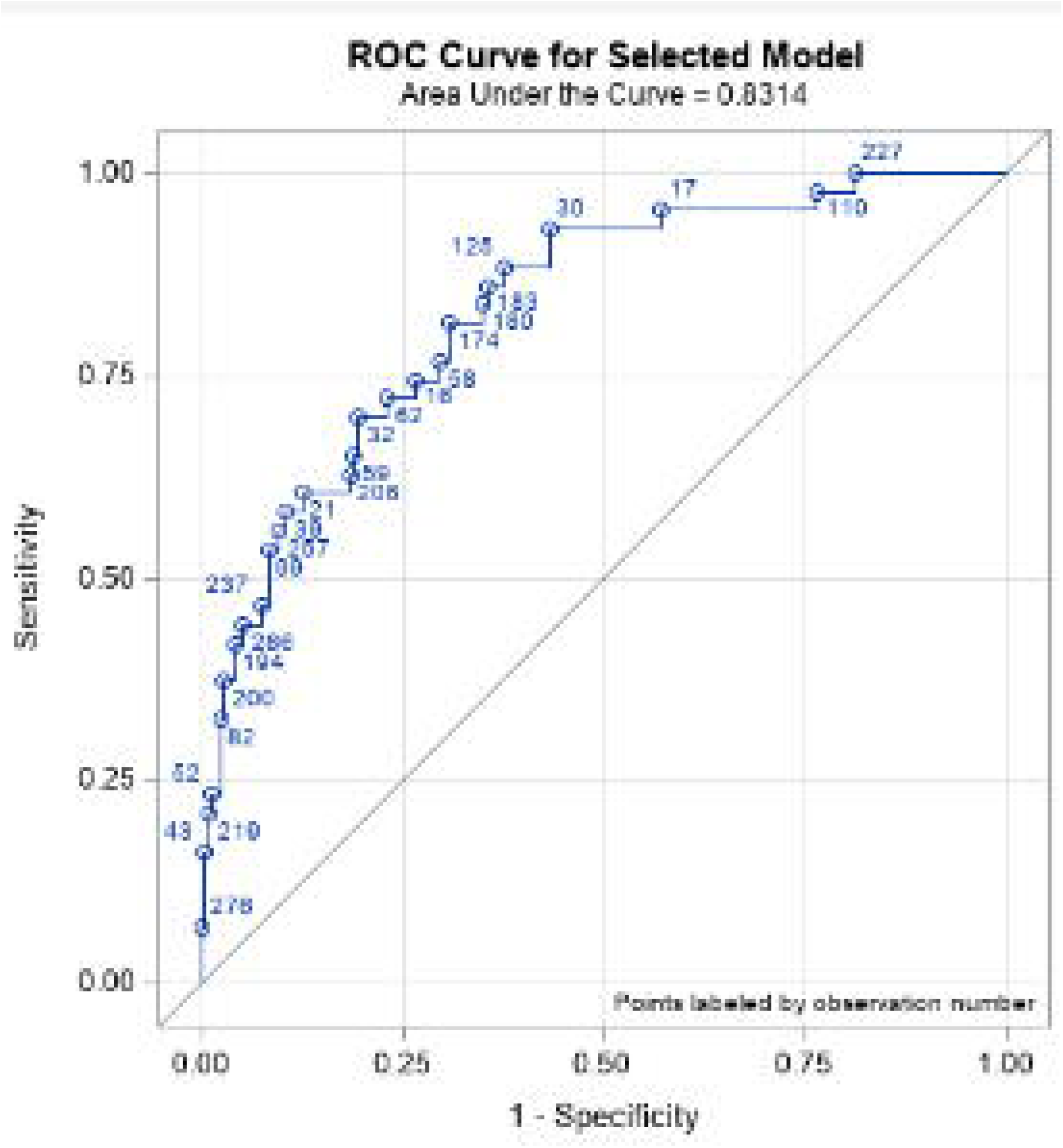
ROC Curve, BMI Sensitivity Analysis

Modeling with a cohort excluding patients with active DNI likewise did not result in meaningful change (AUC = 0.82; Supplemental Figure 4).

The ordinal logit model (Table 4) reached similar parameter selection, but had slightly lower concordance (80%) than the binary model (85%) and did not include admission CRP or biological sex. The proportional odds assumption was not obviously violated by chi square testing (p=0.11).

**Table 4.**
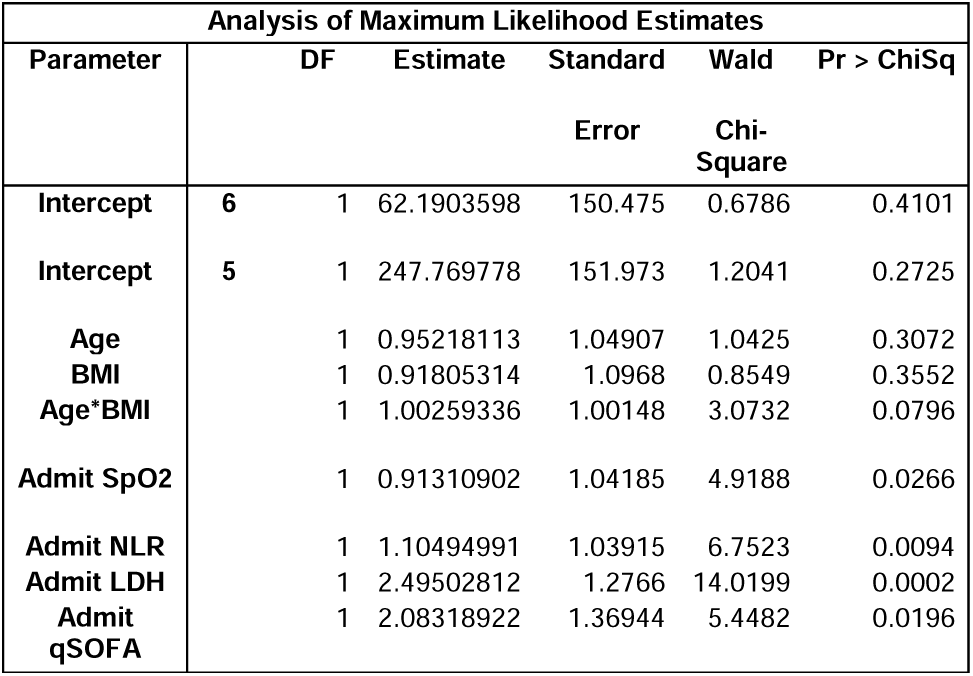
Ordinal Logistic Regression Analysis.

## DISCUSSION

Our study evaluated demographic and clinical variables measured within 12 hours of hospital admission in COVID19 patients as potential predictors of progression to severe respiratory failure. We used the WHO ordinal scale 6-9 to define patients with severe respiratory failure requiring significant life sustaining therapies. Our analysis demonstrated that a combination of routine accessible laboratory tests, vital signs, and demographic variables may yield a useful clinical tool to assess risk of COVID-19 severe respiratory failure among patients admitted to our health system. This model used objective and measurable information available in acute care settings even if the patient is unable to communicate. The model could serve as an effective point of service tool during early admission to assist in clinical management and allocation of resources to unvaccinated persons. The significant factors, like admission LDH, were robust to numerous sensitivity analyses.

LDH was highly predictive of severe disease in our model and was robust to sensitivity analysis. Subjects with abnormal LDH on admission were 2.36 times more likely to progress to severe hypoxemic respiratory failure after controlling for other factors. This finding is supported by prior reports that LDH can predict severity of disease.[4, 5] However, our findings differ from those by Liang et al. showing increased LDH only increase the likelihood of severe disease by 0.2%.[4] A recent meta-analysis by Katzenschlager et al. evaluated the association between LDH levels and admission to the intensive care unit (12 studies) or death (23 studies) in patients with COVID 19. Although LDH levels were statistically higher in those critically ill (pooled difference of medians: 140 U/L (95% CI 81-199) and those who died (pooled difference of medians: 189 U/L (95% CI 155-223)), the modest absolute increase in LDH levels was deemed clinically irrelevant by the authors.[26] The differences between these studies and ours may be explained by the different outcome definitions used. In our system, the use of high flow nasal cannula was not necessarily associated with ICU admission but was included as part of the endpoint. The ordinal logistic regression performed, however, supports that collapsing these two outcomes into a composite outcome yields comparable predictive utility.

NLR has been associated with adverse outcomes in COVID19 patients.[4, 7, 9] Adverse outcomes observed in these studies also included death, which likely accounted for the absolute risk difference in our study.[4, 9] Loannou et al. reported that a ratio higher than 12.7 was associated with a 2.5-fold increase in the odds for mechanical ventilation in COVID19 patients.[7] In our cohort, a higher NLR was associated with modest increases in the odds for reaching ordinal scale 6-9. The contrast between our results and those of Loannou et al. may be related to the inclusion of less severe disease categories in our primary endpoint such as receiving high-flow nasal canula. In our cohort, age and BMI were important predictors of COVID-19 respiratory failure. BMI was positively associated with progression to WHO ordinal score 6-9. Because the average BMI was >30 in our study, meaningful stratification analysis was precluded. While the mean BMI imputation biases towards significance, multiple imputation is not appropriate when data are not missing randomly relative to primary outcome. However, excluding patients who did not have valid BMI data did not meaningfully change our model findings. In addition, the overall AUC did not change in comparison to the original model. Although, the age/BMI composite variable was retained by stepwise AIC reduction after excluding cases without valid BMI data, using a composite variable introduces unneeded complexity to the model. Thus, we ultimately decided to exclude the term to maintain simplicity. These data highlighted the association of BMI with severe COVID-19 and add to previous studies that support this association.[7, 22, 28] A proposed mechanism for this association is the increased work of breathing in patients with high body mass index that impairs their capacity to adjust to changes in lung function leading to earlier non-invasive ventilation or mechanical ventilation.[29] We have chosen our predictors based on clinical practicality and mechanistic plausibility. Several factors – D-Dimer, CRP, Sex – were not significant predictors but augmented the AUC collectively. Thus, it is not surprising that some sensitivity analyses (i.e., ordinal logistic regression) did not retain some or all of these factors. While D-dimer elevations were not found to be a significant predictor for respiratory failure or death in some studies.[4, 8], others found an association with adverse outcomes early in the pandemic.[29, 30] Pulmonary vasculature thrombosis was observed in autopsies of COVID-19 patients, [31] and a recent study has suggested that heparin-based anticoagulation may protect noncritical COVID-19 patients from inpatient death.[32] In our model, D-dimer was not statistically associated with developing the primary endpoint in the multivariable analysis.

A smaller proportion of subjects in our cohort met the WHO ordinal scale 6-9 than subjects in other, early 2020 cohorts. Only 19% of the subjects included in our modeling cohort met the primary endpoint compared to 22-26% reported by other authors.[6, 8] Our cohort was enrolled during a phase of rapidly evolving COVID-19 therapies and management approaches. With improvements in early interventions against virus replication and associated inflammation the number of patients requiring high flow oxygen or mechanical ventilation is expected to change. We also found a lower inpatient death rate compared to reports published around the same period.[8] All the subjects included in our cohort have been discharged at the time of data collection. It is possible that a subpopulation of these subjects was re-admitted and expired after data collection was completed. Our study design limited data collection to the primary subject admission and may have missed mortality that occurred in subsequent encounters. We included patients with DNI and comfort care orders in our cohort. Although, this is a group of subjects that would have not been able to reach all the ordinal scale scores in our endpoint, they would have been eligible for high flow oxygen and vasopressors. The sensitivity analysis that omitted subjects with DNI did not significantly change the predictive fidelity of the model. The inclusion of this sub-population in our cohort likely provided a conservative estimate of the odds of meeting ordinal scales 6-9. Our study has several limitations to acknowledge. Troponins were not included in our model because all subjects with abnormal troponin met the primary outcome. Elevated troponin suggested myocardial injury which can be due to a direct effect from SARS-CoV-2 infection and/or a complication from sepsis and the inflammatory response described in COVID 19. The role of troponin as a predictor of COVID-19 associated mortality has been suggested in other studies.[33, 34] However, larger studies are necessary to evaluate their role in predicting severe COVID-19 respiratory failure. Additionally, our cohort was constructed prior to introduction of COVID-19 vaccination and therapeutic interventions such as dexamethasone or remdesivir.[35] Most importantly, the validation of our prediction model in the rest of our study population and in more recent cohorts after the emergence of new SARS-CoV-2 variants will be critical to assess its real-world clinical utility. Our prediction model could contribute by aiding clinicians who desire point-of-care decision support in early COVID-19 disease.

## CONCLUSION

This study provides a preliminary model for early identification of COVID19 patients at odds of progressing to severe COVID-19 within the first 12 hours of admission. This model will require further validation in larger datasets. Future studies will use this model as a tool for predicting severe COVID-19 disease in resource limited settings where effective vaccines and therapies are still unavailable.

## Data Availability

SAS Code will be published at Dryad, an online repository. TDCJ prisoners will be excluded from this data.

## ABBREVIATIONS

(SARS-CoV-2): Severe acute respiratory syndrome-Coronoavius-2
(COVID-19): Coronaviurs-Disease-19
(WHO): World Health Organization
(CDC): Center of Disease Control and Prevention
(UTMB): University of Texas Medical Branch
(TDCJ): Texas Department of Criminal Justice
(GCS): Glawgow coma scale
(qSOFA): Quick Sequential Organ Failure Assessment
(NLR): Neutrophil to lymphocyte ratio
(LDH): Lactate Dehydrogenase
(CRP): C-reactive protein
(BMI): Body mass index
(IQR): Interquartile range
(SD): Standard deviation
(AIC): Stepwise Akaike Information Criteria
(GOF): Goodness of fit
(ROC): Receiver operating characteristics curve
(AUC): Area under the curve

## CONTRIBUTORS

Fu C and Harmouch W both contributed to data collection and manuscript drafting. Reynoso DR and Keiser P both contributed to project development and data allocation. Cabada MC has been the senior author since project inception and providing editorial revision to the manuscript. Davis JW designed and performed all statistical analysis and drafted the methods, results, and discussion sections in conjunction with Tomczak E and Wang B. Tomczak E was responsible for project conception, some data collection, and some manuscript drafting, and Wang B was responsible for the project conception, data collection execution and drafting all parts of manuscript.

## ACKNOWLEDGMENTS

We thank Daniel Z. Bao, Ashley E Chen and Kyra Curtis from UTMB Medical School for their contribution to data collection. We thank Dr. Ion Mitrache, Director of Clinical Analyst at UTMB, for assistance in clinical data management.

## DATA AVAILABILITY STATEMENT

SAS Code and de-identified data will be published at Dryad, an online repository. TDCJ prisoners will not be included.

## ETHICS STATEMENT

The study protocol has obtained waiver of informed consent under the approvals of Institutional Review Board from UTMB (20-0126) and Texas Department of Criminal Justice (TDCJ) under protocol number #819-RM20.

## REFERENCES

1. WHO. COVID-19 clinical management. Living guidance. 2021 January 25, 2021.[cited 2021 May 13th]; Second:.[Available from: https://www.who.int/publications/i/item/WHO-2019-nCoV-clinical-2021-1.

2. WHO. Past pandemics [cited 2021 June 11th]; Available from: https://www.euro.who.int/en/health-topics/communicable-diseases/influenza/pandemic-influenza/past-pandemics.

3. Wu, Z. and J.M. McGoogan, Characteristics of and Important Lessons From the Coronavirus Disease 2019 (COVID-19) Outbreak in China: Summary of a Report of 72□314 Cases From the Chinese Center for Disease Control and Prevention. Jama, 2020. 323(13): p. 1239–1242.

4. Liang, W., et al., Development and Validation of a Clinical Risk Score to Predict the Occurrence of Critical Illness in Hospitalized Patients With COVID-19. JAMA Intern Med, 2020. 180(8): p. 1081–1089.

5. Poggiali, E., et al., Lactate dehydrogenase and C-reactive protein as predictors of respiratory failure in CoVID-19 patients. Clin Chim Acta, 2020. 509: p. 135–138.

6. Cummings, M.J., et al., Epidemiology, clinical course, and outcomes of critically ill adults with COVID-19 in New York City: a prospective cohort study. Lancet, 2020. 395(10239): p. 1763–1770.

7. Ioannou, G.N., et al., Risk Factors for Hospitalization, Mechanical Ventilation, or Death Among 10□131 US Veterans With SARS-CoV-2 Infection. JAMA Netw Open, 2020. 3(9): p. e2022310.

8. Garibaldi, B.T., et al., Patient Trajectories Among Persons Hospitalized for COVID-19 : A Cohort Study. Ann Intern Med, 2021. 174(1): p. 33–41.

9. Wongvibulsin, S., et al., Development of Severe COVID-19 Adaptive Risk Predictor (SCARP), a Calculator to Predict Severe Disease or Death in Hospitalized Patients With COVID-19. Ann Intern Med, 2021. 174(6): p. 777–785.

10. WHO. WHO R&D Bluprint. Novel Coronavirus. COVID-19 Therapeutic Trial Synoposis. 2020 Februrary 18, 2020.[cited 2021 May 15th]; Available from: https://www.who.int/publications/i/item/covid-19-therapeutic-trial-synopsis

11. Beigel, J.H., et al., Remdesivir for the Treatment of Covid-19 - Final Report. N Engl J Med, 2020. 383(19): p. 1813–1826.

12. Kalil, A.C., et al., Baricitinib plus Remdesivir for Hospitalized Adults with Covid-19. N Engl J Med, 2021. 384(9): p. 795–807.

13. Midi H, Sarkar SK, Rana S. Collinearity diagnostics of binary logistic regression model. J Interdiscip Math Published Online First: 28 May 2013.http://www.tandfonline.com/doi/abs/10.1080/09720502.2010.10700699 (accessed 4 Jan 2022).

14. Ranney, M.L., V. Griffeth, and A.K. Jha, Critical Supply Shortages - The Need for Ventilators and Personal Protective Equipment during the Covid-19 Pandemic. N Engl J Med, 2020. 382(18): p. e41.

15. Gundlapalli, A.V., et al., Death Certificate-Based ICD-10 Diagnosis Codes for COVID-19 Mortality Surveillance - United States, January-December 2020. MMWR Morb Mortal Wkly Rep, 2021. 70(14): p. 523–527.

16. Guy, G.P., Jr., et al., Association of State-Issued Mask Mandates and Allowing On-Premises Restaurant Dining with County-Level COVID-19 Case and Death Growth Rates - United States, March 1-December 31, 2020. MMWR Morb Mortal Wkly Rep, 2021. 70(10): p. 350–354.

17. Christie, A., et al., Decreases in COVID-19 Cases, Emergency Department Visits, Hospital Admissions, and Deaths Among Older Adults Following the Introduction of COVID-19 Vaccine - United States, September 6, 2020-May 1, 2021. MMWR Morb Mortal Wkly Rep, 2021. 70(23): p. 858–864.

18. Joo, H., et al., Decline in COVID-19 Hospitalization Growth Rates Associated with Statewide Mask Mandates - 10 States, March-October 2020. MMWR Morb Mortal Wkly Rep, 2021. 70(6): p. 212–216.

19. CDC. COVID Data Tracker Weekly Review 8/5/2021. 2021 [cited 2021 8/5]; Available from: https://www.cdc.gov/coronavirus/2019-ncov/covid-data/covidview/index.html.

20. Officials, W.H.C.-R.T.a.P.H., Press Briefing by White House COVID-19 Response Team and Public Health Officials. Aug 2, 2021: The White House, United States.

21. Wynants, L., et al., Prediction models for diagnosis and prognosis of covid-19: systematic review and critical appraisal. Bmj, 2020. 369: p. m1328.

22. Simonnet, A., et al., High Prevalence of Obesity in Severe Acute Respiratory Syndrome Coronavirus-2 (SARS-CoV-2) Requiring Invasive Mechanical Ventilation. Obesity (Silver Spring), 2020. 28(7): p. 1195–1199.

23. Tjendra, Y., et al., Predicting Disease Severity and Outcome in COVID-19 Patients: A Review of Multiple Biomarkers. Arch Pathol Lab Med, 2020. 144(12): p. 1465–1474.

24. Wu, C., et al., Risk Factors Associated With Acute Respiratory Distress Syndrome and Death in Patients With Coronavirus Disease 2019 Pneumonia in Wuhan, China. JAMA Intern Med, 2020. 180(7): p. 934–943.

25. Ji, D., et al., Prediction for Progression Risk in Patients With COVID-19 Pneumonia: The CALL Score. Clin Infect Dis, 2020. 71(6): p. 1393–1399.

26. A minimal common outcome measure set for COVID-19 clinical research. Lancet Infect Dis, 2020. 20(8): p. e192–e197.

27. Katzenschlager, S., et al., Can we predict the severe course of COVID-19 - a systematic review and meta-analysis of indicators of clinical outcome? PLoS One, 2021. 16(7): p. e0255154.

28. Anderson, M.R., et al., Body Mass Index and Risk for Intubation or Death in SARS-CoV-2 Infection : A Retrospective Cohort Study. Ann Intern Med, 2020. 173(10): p. 782–790.

29. Liao, D., et al., Haematological characteristics and risk factors in the classification and prognosis evaluation of COVID-19: a retrospective cohort study. Lancet Haematol, 2020. 7(9): p. e671–e678.

30. Zhou, F., et al., Clinical course and risk factors for mortality of adult inpatients with COVID-19 in Wuhan, China: a retrospective cohort study. Lancet, 2020. 395(10229): p. 1054–1062.

31. Ackermann, M., et al., Pulmonary Vascular Endothelialitis, Thrombosis, and Angiogenesis in Covid-19. N Engl J Med, 2020. 383(2): p. 120–128.

32. Chowdhury, J.F., L.K. Moores, and J.M. Connors, Anticoagulation in Hospitalized Patients with Covid-19. N Engl J Med, 2020. 383(17): p. 1675–1678.

33. García de Guadiana-Romualdo, L., et al., Cardiac troponin and COVID-19 severity: Results from BIOCOVID study. Eur J Clin Invest, 2021. 51(6): p. e13532.

34. Wibowo, A., et al., Prognostic performance of troponin in COVID-19: A diagnostic meta-analysis and meta-regression. Int J Infect Dis, 2021. 105: p. 312–318.

35. Coronavirus Disease 2019 (COVID-19) Treatment Guidelines. Aug 17th 2021 August 17, 2021.[cited 2021 August 20, 2021]; Available from: https://www.covid19treatmentguidelines.nih.gov/about-the-guidelines/whats-new/.

