## Supplement for "Prediction of the need for intensive oxygen supplementation during the hospitalization among patients with COVID 19 admitted to an academic health system in Texas, USA: a retrospective cohort study and development of a multivariable regression model"

Supplemental **Table A. Variable of Interest List**

| | |
|---|---|
| **Demographics** | Date of Birth<br>Sex<br>Body Mass Index (BMI)<br>Admission Campus |
| **Clinical data** | Discharge status<br>Discharge date<br>Date of death<br>Do not intubate (DNI) status<br>First Vitals<br>Presenting symptoms<br>Symptom duration<br>Comorbidities (diabetes mellitus, cardiovascular, pulmonary, and kidney disease)<br>Oxygen supplementation modality (if any)<br>Oxygen flow requirements.<br>Glasgow coma scale (GCS) < 15 vs = 15 (disorientation or other indication from clinical note)<br>Systolic blood pressure, Respiratory Rate, and GCS were used to calculate the Quick Sequential Organ Failure Assessment (qSOFA). |
| **Comorbidities** | Cardiovascular diseases included:<br>Hypertension<br>Coronary artery disease<br>Heart failure<br>History of cardiac resynchronization device placement or left ventricular assisting device placement |
| | Lung diseases included:<br>Asthma<br>Chronic Obstructive Airway Disease<br>Pulmonary hypertension<br>Obstructive sleep apnea<br>Interstitial lung disease<br>Use of home oxygen<br>Use of home continuous positive airway pressure (CPAP) or bilevel positive airway pressure (BiPAP). |
| | Chronic kidney disease included:<br>Any stage 1-5 or end stage renal disease requiring renal replacement therapy. |

Supplemental Figure 1. Original cohort characteristics prior to exclusion, N = 352

| | WHO ordinal scores <6 N =287 | WHO ordinal scores 6-9 N =65 | |
|---|---|---|---|
| | N (%) | | p- values |
| Symptoms | | | |
| Cough | 147 (51.2) | 39 (60.0) | - |
| Dyspnea | 150 (52.3) | 45 (69.2) | |
| Rhinorrhea | 13 (4.5) | 5 (7.7) | |
| Anosmia | 16 (5.6) | 3 (4.6) | |
| Dysgeusia or hypogeusia | 27 (9.4%) | 9 (13.8) | |
| Sore throat | 15 (5.2) | 4 (6.2) | |
| Fever | 116 (40.4) | 36 (55.4) | |
| Hemoptysis | 4 (1.4) | 0 (0) | |
| Nausea and/or vomiting | 66 (23) | 10 (15.4) | |
| Diarrhea | 54 (18.8) | 20 (30.8) | |
| Myalgia and/or arthritis | 47 (16.4) | 11 (16.9) | |
| Rash | 1 (0.3) | 0 (0) | |
| Altered mental status | 16 (5.6) | 5 (7.7) | |
| Asymptomatic | 54 (18.8) | 4 (6.2) | |
| Comorbidities | | | |
| Cardiovascular | 172 (59.9) | 41 (63.1) | - |
| Diabetes mellitus | 87 (30.3) | 24 (36.9) | |
| Lung disease | 56 (19.5) | 8 (12.3) | |
| Renal | 34 (11.8) | 6 (9.2) | |
| Liver | 23 (8) | 10 (15.4) | |
| HIV | 2 (0.7) | 2 (3.1) | |
| Malignancy on chemotherapy | 4 (1.4) | 1 (1.5) | |
| Solid organ transplant | 6 (2.1) | 1 (1.5) | |
| Dementia | 16 (5.6) | 3 (4.6) | |
| Others | 46 (16.0) | 6 (9.2) | |
| No known PMH | 35 (12.2) | 8 (12.3) | |
| Campuses | | | |
| Angleton | 22 (7.7) | 4 (6.2) | - |
| League City | 44 (15.3) | 17 (26.2) | |

| | | | |
|---|---|---|---|
| Clear Lake | 51 (17.8) | 9 (13.8) | |
| Galveston | 165 (57.5) | 32 (49.2) | |
| Non UTMB transfers | 4 (1.4) | 2 (3.1) | |
| Missing values | 1 (0.3) | 1 (1.5) | |
| Demographics | | | |
| Sex | | | |
| Male | 165 (57.5) | 44 (67.7) | 0.17 |
| Female | 122 (14.5) | 21 (32.3) | |
| Code status | | | |
| Comfort care | 8 (2.8) | 10 (15.4) | - |
| DNI | 18 (6.3) | 6 (9.2) | |
| Full code | 261 (90.9) | 47 (72.3) | |
| Missing values | 0 (0) | 2 (3.1) | |
| **Clinical variables **** | Mean (± SD) | | p- values |
| BMI | 31.4 (7.43) | 33.9 (8.86) | 0.0408 |
| O2 saturation, % | 95.6 (6.58) | 93.0 (5.62) | 0.00133 |
| NLR | 5.69 (5.10) | 9.92 (11.2) | 0.00505 |
| CRP | 4.46 (7.62) | 11.5 (10.2) | < 0.001 |
| D-dimer | 1.99 (7.86) | 3.62 (16.6) | 0.449 |
| qSOFA | 0.286 (0.475) | 0.569 (0.558) | <0.001 |
| LDH | 402 (418) | 783 (559) | <0.001 |
| | N (%) | | p- values |
| Categorical LDH | | | |
| Normal value | 197 (68.6) | 19 (29.2) | <0.001 |
| 1-2 x ULN | 77 (2638) | 30 (46.2) | |
| > 2 x ULN | 13 (4.5) | 14 (21.5) | |

** 2 subjects in ordinal score 6-9 group had consistently missed values on clinical variables

Supplemental Figure 2. Initial regression model of cohort, N=329

| Analysis of Maximum Likelihood Estimates | | | | | |
|---|---|---|---|---|---|
| **Parameter** | **DF** | **Estimate** | **Standard Error** | **Wald Chi-Square** | **Pr > Chi Sq** |
| **Intercept** | 1 | 3.18E+55 | 1.97092E+47 | 1.3761 | 0.2408 |
| **Sex** | 1 | 0.918237 | 6.234510078 | 0.0022 | 0.9628 |
| **Age** | 1 | 0.985309 | 1.066838925 | 0.0521 | 0.8195 |
| **Sex*Age** | 1 | 0.991982 | 1.028910011 | 0.0797 | 0.7777 |
| **Admission Date** | 1 | 0.994326 | 1.004952222 | 1.3234 | 0.25 |
| **BMI** | 1 | 0.945066 | 1.119743902 | 0.2499 | 0.6171 |
| **Age*BMI** | 1 | 1.002062 | 1.001881768 | 1.2095 | 0.2714 |
| **Admission SpO2** | 1 | 0.930345 | 1.052638638 | 1.9798 | 0.1594 |
| **Duration Sx** | 1 | 0.947432 | 1.106276642 | 0.2857 | 0.593 |
| **Admit NLR** | 1 | 1.178686 | 1.064281581 | 6.9766 | 0.0083 |
| **Admit Procalcitonin** | 1 | 0.835939 | 1.566587991 | 0.1594 | 0.6897 |
| **Admit D-Dimer** | 1 | 0.938943 | 1.046341715 | 1.9368 | 0.164 |
| **Admit LDH** | 1 | 2.308505 | 1.372179017 | 6.993 | 0.0082 |
| **Admit CRP** | 1 | 1.807058 | 1.546354152 | 1.8428 | 0.1746 |
| **qSOFA** | 1 | 2.932338 | 1.429178781 | 9.0749 | 0.0026 |

*Note that complete case exclusion not applied for this table

Supplemental Figure 3. **Stratified BMI Analysis**

| Analysis of Maximum Likelihood Estimates | | | | | |
|---|---|---|---|---|---|
| **Parameter** | **DF** | **Estimate** | **Standard Error** | **Wald Chi-Square** | **Pr > Chi Sq** |
| **Intercept** | 1 | 0.000554 | 6.30599 | 16.5798 | <.0001 |
| **Age** | 1 | 1.032931 | 1.019284 | 2.8751 | 0.09 |
| **Admit NLR** | 1 | 1.190294 | 1.09319 | 3.8177 | 0.0507 |
| **Admit LDH** | 1 | 5.452641 | 1.594085 | 13.2276 | 0.0003 |

**Lower BMI Analysis (BMI <30)**

| Analysis of Maximum Likelihood Estimates | | | | | |
|---|---|---|---|---|---|
| **Parameter** | **DF** | **Estimate** | **Standard Error** | **Wald Chi-Square** | **Pr > Chi Sq** |
| **Intercept** | 1 | 7.73534434 | 204.95696 | 0.1477 | 0.7007 |
| **Sex** | 1 | 0.32184024 | 1.6594729 | 5.0094 | 0.0252 |
| **Age** | 1 | 1.05812661 | 1.0196914 | 8.401 | 0.0038 |
| **BMI** | 1 | 1.06822672 | 1.0338607 | 3.9238 | 0.0476 |
| **Admit SpO2** | 1 | 0.90429468 | 1.054746 | 3.5653 | 0.059 |
| **Admit NLR** | 1 | 1.16602438 | 1.0778842 | 4.1962 | 0.0405 |
| **Admit D-Dimer** | 1 | 0.65422758 | 1.215311 | 4.736 | 0.0295 |
| **Admit LDH** | 1 | 2.40873087 | 1.4663847 | 5.2744 | 0.0216 |
| **Admit CRP** | 1 | 2.86967849 | 1.708473 | 3.8735 | 0.0491 |

**Higher BMI Analysis (BMI => 30)**

Supplemental Figure 4. DNI Sensitivity Analysis

| Analysis of Maximum Likelihood Estimates | | | | | |
|---|---|---|---|---|---|
| Parameter | DF | Estimate | Standard Error | Wald Chi-Square | Pr > Chi Sq |
| Intercept | 1 | 77.2154835 | 175.3002 | 0.7078 | 0.4002 |
| Age | 1 | 0.97384804 | 1.051061 | 0.2828 | 0.5949 |
| BMI | 1 | 0.93304672 | 1.104839 | 0.4836 | 0.4868 |
| Age*BMI | 1 | 1.00215231 | 1.001561 | 1.8954 | 0.1686 |
| Admit SpO2 | 1 | 0.92154838 | 1.043312 | 3.7208 | 0.0537 |
| Admit NLR | 1 | 1.07422557 | 1.026238 | 7.66 | 0.0056 |
| Admit D-Dimer | 1 | 0.98049278 | 1.01349 | 2.1498 | 0.1426 |
| Admit LDH | 1 | 3.46668914 | 1.288785 | 24.006 | <.0001 |
| qSOFA | 1 | 2.37049786 | 1.370533 | 7.5001 | 0.0062 |
